# Initial SARS-CoV-2 vaccination response can predict booster response for BNT162b2 but not for AZD1222

**DOI:** 10.1101/2021.07.06.21260059

**Authors:** Thomas Perkmann, Nicole Perkmann-Nagele, Patrick Mucher, Astrid Radakovics, Manuela Repl, Thomas Koller, Galateja Jordakieva, Oswald F Wagner, Christoph J Binder, Helmuth Haslacher

## Abstract

**Objectives:** Our objective was to determine whether SARS-CoV-2 antibody levels after the first dose can predict the final antibody response and whether this is dependent on the vaccine type.

**Methods:** 69 BNT162b2 (Pfizer/BioNTech) and 55 AZD1222 (AstraZeneca) vaccinees without previous infection or immunosuppressive medication were included. Anti-body levels were quantified 3 weeks after dose 1, in case of AZD1222 directly before boostering (11 weeks after dose 1) and 3 weeks after dose 2, with the Roche SARS-CoV-2 S total antibody assay.

**Results:** Pre-booster (BNT162b2: 80.6 [25.5-167.0] BAU/mL, AZD1222: 56.4 [36.4-104.8] BAU/mL, not significant) and post-booster levels (BNT162b2: 2,092.0 [1,216.3-4,431.8] BAU/mL, AZD1222: 957.0 [684.5-1,684.8] BAU/mL, p<0.0001) correlated well in BNT162b2 (ρ=0.53) but not in AZD1222 recipients. Moreover, antibody levels after the first dose of BNT162b2 correlated inversely with age (ρ=-0.33, P=0.013), whereas a positive correlation with age was observed after the second dose in AZD1222 recipients (ρ=0.26, P=0.030).

**Conclusions:** In conclusion, our data suggest that antibody levels quantified by the Roche Elecsys SARS-CoV-2 S assay before the booster shot could infer post-booster responses to BNT162b2, but not to AZ1222. In addition, we found a vaccine-dependent effect on antibody responses, suggesting a possible link between vaccine response and vector immunity.

## Background

Vaccines against the novel coronavirus SARS-CoV-2 have now been available for several months (Zaqout et al., 2021). Determination of spike-protein specific antibodies after SARS-CoV-2 vaccination, although not unrestrictedly recommended (Centers for Disease Control and Prevention, 2021), is commonly performed. The post-vaccination antibody levels, even when measured with standardized commercially available CE-certified quantitative test systems, differ significantly (Kristiansen et al., 2021, Perkmann et al., 2021a). Furthermore, in addition to these analytically related differences, there are significant differences in expected levels depending on the age and serostatus of the vaccinees (Krammer et al., 2021, Perkmann et al., 2021b, Subbarao et al., 2021), the vaccine used (Eyre et al., 2021), and the timing of blood collection (elapsed time interval since first or second dose).

To date, it is not clear to what extent antibody levels after the first dose are suitable to infer the booster response. Similarly, it is unclear whether this response depends on the type of vaccine used. Indeed, this would be likely because vector and mRNA vaccines elicit different immune responses, with vector vaccines also including a non-spike-specific response directed against the vector (Federico, 2021).

We report here differences in the predictability of SARS-CoV-2 vaccine post-booster levels measured with a quantitative antibody assay (Roche Elecsys® SARS-CoV-2 S) that depend on the vaccine used. We also present the differential impact of age on the antibody response to AZD1222 (AZD1222, Astra Zeneca) or BNT162b2 (BNT162b2, Pfizer/BioNTech).

## Methods

Of 166 participants recruited within the MedUni Wien Biobank’s healthy donors’ collection until March 5^th^ 2021, 124 were eligible for inclusion. All subjects were >18 years old and provided written informed consent to participate in the study. Reasons for exclusion were a previous infection with SARS-CoV-2 and ongoing immunosuppressive medication, as these conditions are known to bias the average vaccination response. In addition, dropouts due to a missed blood sampling and the onset of COVID between the first and second doses occurred (see Figure 1). The prime-boost regimen specified an 11-week dosing interval for AZD1222 and a 3-week dosing interval for BNT162b2. The protocol of this performance evaluation study was reviewed and approved by the Ethics committee of the Medical University of Vienna (EK 1066/2021).

**Fig. 1.**
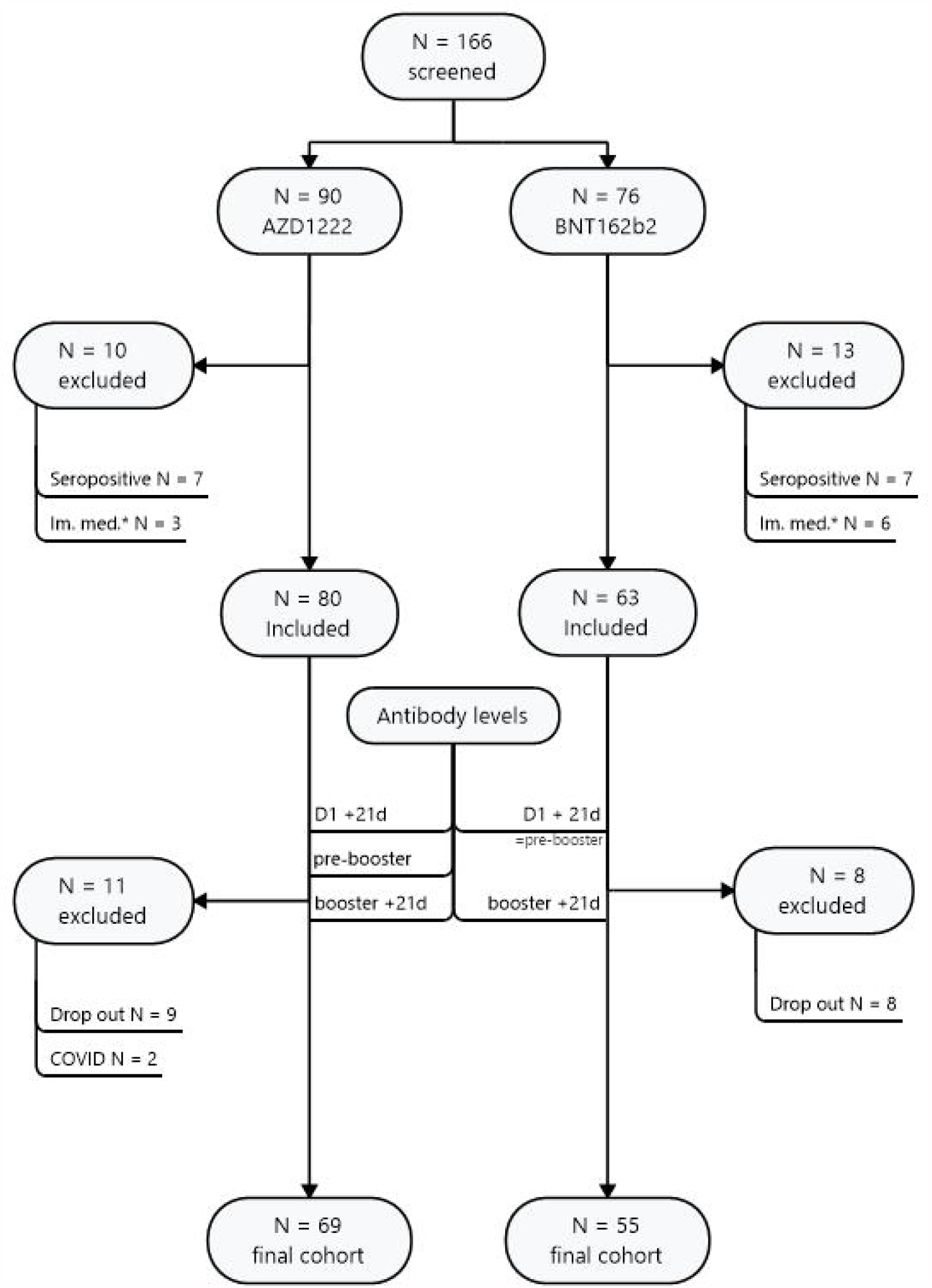
Study flow chart

Samples were processed and, if applicable, stored before analysis at <-70°C according to standard operating procedures by the MedUni Wien Biobank in an ISO 9001:2015-certified environment (Haslacher et al., 2018). A previous SARS-CoV-2 infection was ruled out or confirmed by the Roche Elecsys SARS-CoV-2 anti-Nucleocapsid total antibody ECLIA (electrochemiluminescence assay) and assumed in all participants with a PCR-proven SARS-CoV-2 infection. Vaccine-induced anti-spike antibodies were quantified using the Roche Elecsys® SARS-CoV-2 S total antibody ECLIA on Roche cobas® e801 modular analyzers. All analyses were performed at the Department of Laboratory Medicine, Medical University of Vienna, which operates a certified (ISO 9001:2015) and accredited (ISO 15189:2012) quality management system. Performance data of both tests have been published before (Perkmann et al., 2020, Perkmann et al., 2021a).

Continuous data, given as median (interquartile range), were compared by rank sign tests (Mann Whitney U test, Wilcoxon test). Categorical data, presented as counts and percentages, were compared by χ²-tests. Rank correlations were computed according to Pearson and presented by Pearson’s ρ. P-values <0.05 were considered statistically significant. All calculations were performed with MedCalc 19.7 (MedCalc, Ostend, Belgium), figures were drawn with Mindjet Manager 19 (Corel, Ottawa, Canada) and Prism 9 (GraphPad, La Jolla, USA).

## Results

### Pre-booster antibody levels predict BNT162b2, but not AZD1222 post-booster levels

The 69 individuals receiving AZD1222 did not significantly differ from the 55 participants vaccinated with BNT162b2 in terms of age (42 [29-50] years vs. 42 [30-53.5] years, P=0.387). However, the proportion of females was higher among the AZD1222 recipients (57/69 [83%] vs. 31/55 [56%], χ²=10.1, P=0.001). Pre-booster levels (at 11 weeks and 3 weeks after first dose, respectively) were 56.4 (36.4-104.8) BAU/mL for AZD1222 and 80.6 (25.5-167.0) BAU/mL for BNT162b2 (P=0.513). 21 (21-22) days after the booster shot, antibody levels rose to 957.0 (684.5-1,684.8) BAU/mL in AZD1222 and to 2,092.0 (1,216.3-4,431.8) BAU/mL in BNT162b2 recipients (P<0.0001).

Correlation of SARS-CoV-2 antibody levels before the booster shot and the antibody response assessed 21 (21-22) days after the booster was calculated. The correlation was significant for Corminaty recipients (ρ=0.53, P<0.0001), but not for those receiving AZD1222 (ρ=0.10, P=0.393), see Fig. 2.

**Fig. 2:**
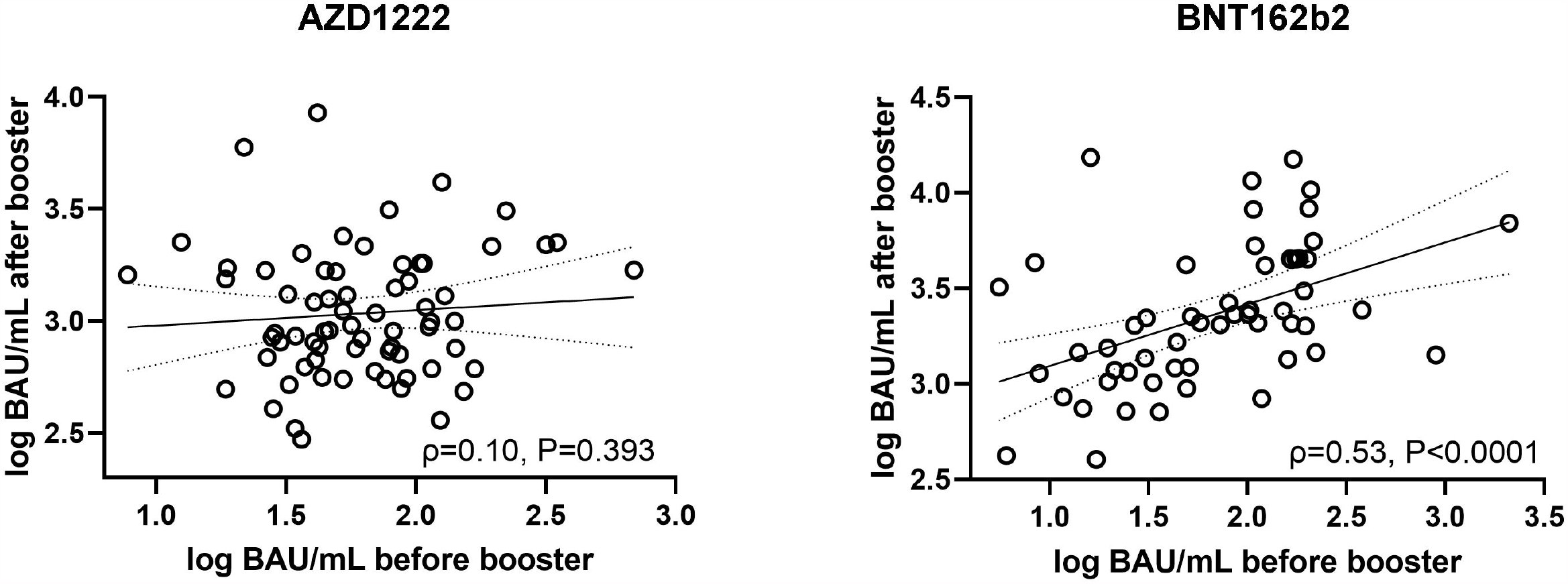
Correlation (according to Spearman) between pre- and post-booster antibody levels (Roche Elecsys S total antibody ECLIA) in AZD1222 (left) and BNT162b2 (right) vaccinees. BAU/mL… binding antibody units per milliliter.

### Antibody levels increase steadily between 3 and 12 weeks after AZD1222

In the next step, the changes of SARS-CoV-2 antibody levels between 3 and 11 weeks after the first AZD1222 dose were assessed. Levels were significantly higher before the booster shot (11 weeks) compated to three weeks after dose 1 (56.4 [36.4-104.8] BAU/mL vs. 13.4 [5.2-27.8] BAU/mL, P<0.0001, see Figure 3A). Levels of both time points correlated at ρ=0.45 (Figure 3B). As for the pre-booster levels, the levels assessed three weeks after dose 1 did not significantly correlate with antibody responses to the booster shot (ρ=0.20, P=0.101).

**Fig. 3:**
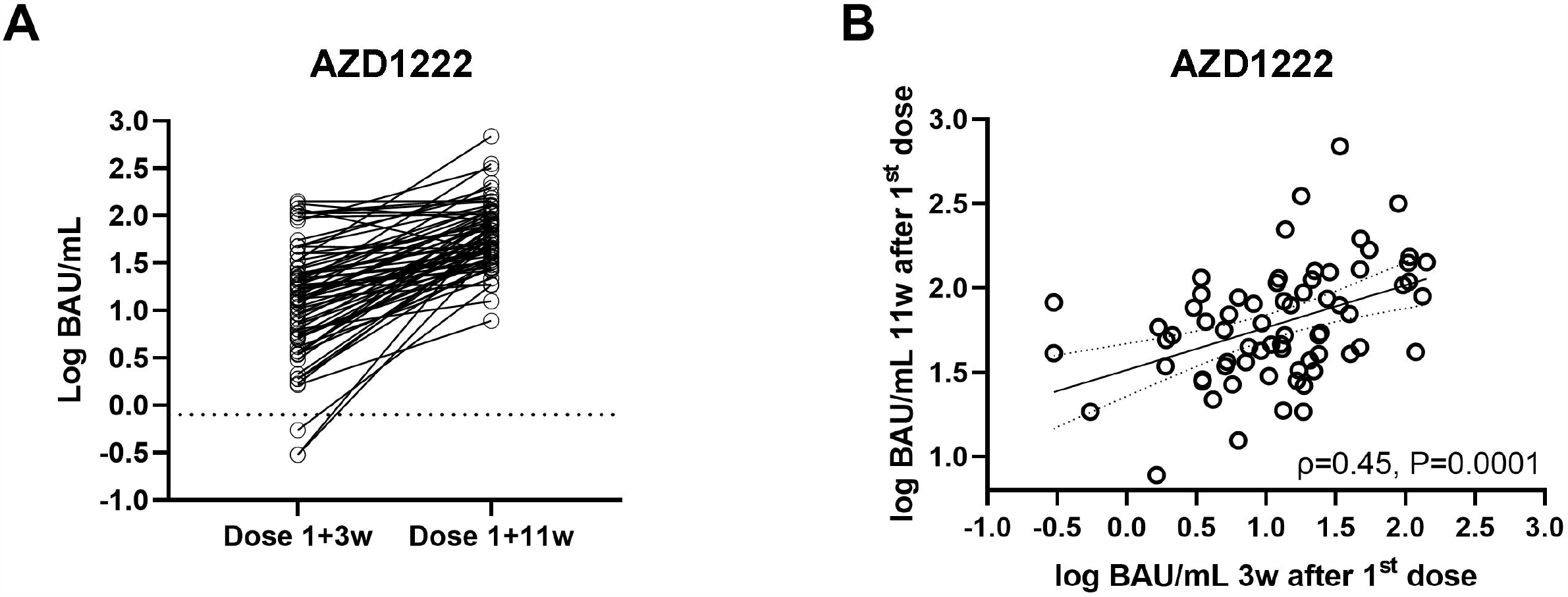
A) Intraindividual progression (A) and correlation (B) of antibody levels (Roche Elecsys S total antibody ECLIA) after AZD1222, measured 3 weeks after first vaccination and before booster (11 weeks). BAU/mL… binding antibody units per milliliter.

### AZD1222 and BNT162b2 behave in opposite ways regarding the correlation of age and antibody response

Finally, we wanted to determine whether the age of the participants affected the antibody responses to the two vaccines. For AZD1222, age did not correlate with the antibody levels measured three weeks after dose 1 (ρ=-0.06, P=0.637) or just before dose 2 (ρ=-0.01, P=0.909). However, there was a weak positive association between age and the antibody response to dose 2 (ρ=0.26, P=0.030). In contrast, antibody levels assessed three weeks after the first Corminaty dose correlated inversely with age (ρ=-0.33, P=0.013); however, this correlation was mitigated after dose 2 (ρ=-0.23, P=0.093), see Figure 4.

**Fig. 4:**
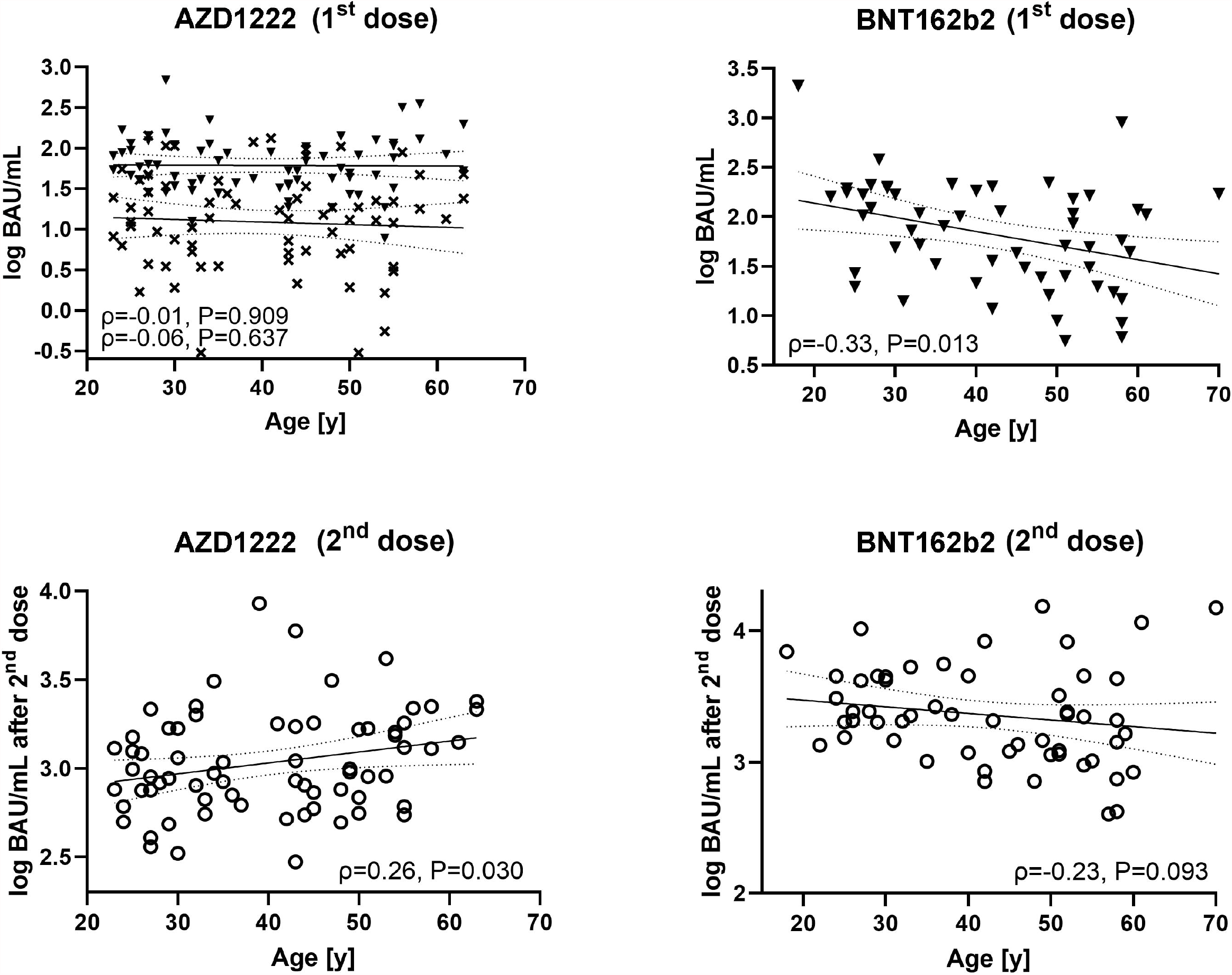
Correlation (according to Spearman) between antibody levels (Roche Elecsys S total antibody ECLIA) and age for AZD1222 (left column) and BNT162b2 (right column) at different points in time (x… 3 weeks after dose 1 in AZD1222, filled triangles… pre-booster, top row; open circles… post-booster, bottom row) BAU/mL… binding antibody units per milliliter).

## Discussion

The relationship between the detection of SARS-CoV-2 specific antibodies and immunity is becoming increasingly well-established based on evidence from numerous studies (Hall et al., 2021, Lumley et al., 2021). Thus, identifying neutralizing antibodies to SARS-CoV-2 as a surrogate marker for established immunity is under discussion (Feng et al., 2021). In this sense, the association with neutralization assays has already been investigated for several SARS-CoV-2 antibody-binding assays (Bal et al., 2021, Perkmann et al.). It has also been shown for the Roche S assay that antibody levels >15 BAU/ml strongly correlate with the presence of neutralizing antibodies (see product manual V2.0). However, the interpretation of antibody levels after vaccination is limited due to the many variables involved. For example, whether initially weak antibody responses also lead to lower levels after the booster and whether this varies with different vaccines has been unclear to date. Therefore, additional data are needed to inform different health policies and clinical decisions (European Centre for Disease Prevention and Control, 2021).

Our data strongly emphasize that the first dose antibody response can predict the final post-booster antibody levels using the mRNA vaccine BNT162b2. However, we were unable to show this relationship for the vector vaccine AZD1222. To the best of our knowledge, this finding was not reported before and might be explained by the specific immune response to vector vaccines. In contrast to mRNA-based preparations, vector vaccines also induce an immune response directed against the viral vector, which in the case of AZD1222 is the chimpanzee adenovirus ChAdOx1 (Kaur and Gupta, 2020). De-novo anti-vector immunity could affect spike-protein induction and, therefore, strongly influence the association between pre- and post-booster values. Whether this demonstrated difference between BNT162b and AZD1222 is transferable to other mRNA and vector vaccines remains to be investigated by further studies.

Above that, we found significant associations between antibody levels and age. With AZD1222, neither of the two pre-booster samples (3 weeks and 11 weeks after dose 1) correlated with age. However, after the booster shot, we found a significant positive association (ρ=-0.26, P=0.013), indicating that older age was associated with slightly increasing antibody levels. A possible explanation might be reduced anti-vector-immunity in older individuals (Connors et al., 2021, Dorrington and Bowdish, 2013).

For BNT162b2, in contrast, we found an inverse correlation of antibody levels with age after dose 1 (ρ=-0.33, P=0.013). However, this association nearly vanished after dose 2 and lost statistical significance (ρ=-0.23, P=0.093). These findings align with recent articles, which found a gap in antibody levels of younger and older individuals after the dose 1 (Müller et al., 2021, Viana et al., 2021).

In conclusion, our data suggest that antibody levels quantified by the Roche Elecsys SARS-CoV-2 S assay before the booster shot could infer post-booster responses to BNT162b2, but not to AZD1222. In addition, we found a vaccine-dependent effect on antibody responses, suggesting a possible link between vaccine response and vector immunity.

## Data Availability

Data is available for interested researchers upon request in compliance with applicable data protection laws.

## Acknowledgments

We thank all sample donors for their valuable contribution. We further thank Radovanovic-Petrova, Monika Martiny, Jadwiga Konarski, Bernhard Haunold, Maedeh Iravany, and Shohreh Lashgari for perfect technical and administrative assistance.

The Department of Laboratory Medicine received compensations for advertisement on scientific symposia from Roche, and holds a grant for evaluating an in vitro diagnostic device from Roche.

The MedUni Wien Biobank is part of the Austrian biobanking consortium BBMRI.at. There was no additional funding received for the present work.

